# *EventEpi*–A Natural Language Processing Framework for Event-Based Surveillance

**DOI:** 10.1101/19006395

**Authors:** Auss Abbood, Alexander Ullrich, Rüdiger Busche, Stéphane Ghozzi

## Abstract

According to the World Health Organization (WHO), around 60% of all outbreaks are detected using informal sources. In many public health institutes, including the WHO and the Robert Koch Institute (RKI), dedicated groups of epidemiologists sift through numerous articles and newsletters to detect relevant events. This media screening is one important part of event-based surveillance (EBS). Reading the articles, discussing their relevance, and putting key information into a database is a time-consuming process. To support EBS, but also to gain insights into what makes an article and the event it describes relevant, we developed a natural-language-processing framework for automated information extraction and relevance scoring. First, we scraped relevant sources for EBS as done at RKI (WHO Disease Outbreak News and ProMED) and automatically extracted the articles’ key data: **disease, country, date**, and **confirmed-case count**. For this, we performed named entity recognition in two steps: EpiTator, an open-source epidemiological annotation tool, suggested many different possibilities for each. We trained a naive Bayes classifier to find the single most likely one using RKI’s EBS database as labels. Then, for relevance scoring, we defined two classes to which any article might belong: The article is *relevant* if it is in the EBS database and *irrelevant* otherwise. We compared the performance of different classifiers, using document and word embeddings. Two of the tested algorithms stood out: The multilayer perceptron performed best overall, with a precision of 0.19, recall of 0.50, specificity of 0.89, F1 of 0.28, and the highest tested index balanced accuracy of 0.46. The support-vector machine, on the other hand, had the highest recall (0.88) which can be of higher interest for epidemiologists. Finally, we integrated these functionalities into a web application called *EventEpi* where relevant sources are automatically analyzed and put into a database. The user can also provide any URL or text, that will be analyzed in the same way and added to the database. Each of these steps could be improved, in particular with larger labeled datasets and fine-tuning of the learning algorithms. The overall framework, however, works already well and can be used in production, promising improvements in EBS. The source code is publicly available at https://github.com/aauss/EventEpi.

## 1 Introduction

### 1.1 Event-based surveillance

The overall goal in **infectious disease epidemiology** is the detection and subsequent containment of infectious disease outbreaks to minimize health consequences and the burden to the public health apparatus. Surveillance systems are an essential part of efficient early-warning mechanisms [1, 2].

A traditional reporting system facilitates trustworthy and health-based formal sources for epidemiological surveillance [2].The acquisition of this data is mostly a passive process and follows routines established by the legislator and the public health institutes. This process is called **indicator-based surveillance**.

Hints of an outbreak, however, can also be detected through changed circumstances that are known to favor outbreaks, e.g., warm weather might contribute to more salmonellosis outbreaks [3] or a loss of proper sanitation might lead to cholera outbreaks [4]. Therefore, besides traditional surveillance that typically relies on laboratory confirmation, secondary data such as weather, attendance monitoring at schools and workplaces, social media, and the web are also significant sources of information [2].

The monitoring of information generated outside the traditional reporting system and its analysis is called **event-based surveillance** (EBS). EBS can greatly reduce the delay between the occurrence and the detection of an event compared to IBS. It enables epidemiologists to detect and report events before the recognition of human cases in the reporting system of the public health system [2]. Especially on the web, the topicality and quantity of data can be useful to detect even rumors of suspected outbreaks. As a result, more than 60% of the initial outbreak reports refer to such informal sources [5].

Filtering this massive amount of data poses the difficulty of finding the right criteria for which information to consider and which to discard. This task is particularly difficult because it is important that the filter does not miss any important events (sensitivity) while being confident what to exclude (precision). Without such a filter process, it is infeasible to perform EBS on larger data sources. Algorithms in the field of natural language processing (NLP) are well suited to tap these informal resources and help to structure and filter this information automatically and systematically [6].

### 1.2 Motivation and contribution

At RKI, the **The Information Centre for International Health Protection** (*Informationsstelle für Internationalen Gesundheitsschutz*, **INIG**), among other units, performs EBS to identify events relevant to public health in Germany. Their routine tasks are defined in standard operating procedures (SOPs) and include reading online articles from a defined set of sources, evaluating them for relevance, and then manually filling a spreadsheet with information from the relevant articles. This spreadsheet is RKI’s EBS database, called **Incident Database** (*Ereignisdatenbank*, **IDB**). The existence of SOPs and the amount of time spend with manual information extraction and especially data entry lead to the idea to automate parts of this process.

Applying methods of natural language processing and machine learning to the IDB, we developed a pipeline that:

- automatically extracts key entities (disease, country, confirmed-case count, and date of the case count) from an epidemiological article and puts them in a database, making tedious data entry unnecessary;
- scores articles for relevance to allow the most important ones to be shown first;
- provides the results in a web service named *EventEpi* that can be integrated in EBS workflows.

### 1.3 Related work

The Global Rapid Identification Tool System^1^ (GRITS) by the EcoHealth Alliance is a web service that provides automatic analyses of epidemiological texts. It uses EpiTator^2^ to extract crucial information about a text, such as dates or countries mentioned, and suggests the most likely disease it is about. However GRITS cannot be automated and is not customizable. To use it in EBS, one would need to manually copy-paste both URLs and output of the analysis. Furthermore, it does not filter news but only extracts entities as they occur in the text independent of the relevance of the entities.

The *recent disease incidents* page of MEDISYS^3^, which channels PULS^4^, tabularly presents automatically-extracted outbreak information from a vast amount of news sources. However, it is not clear how articles are filtered, how information is extracted, or how uncertain the output is. Therefore, it cannot be used as such and as it is a closed software we could not develop it further.

## 2 Methods

The approach presented here consists of two largely independent, but complementary parts: key information extraction and relevance scoring. Both approaches are integrated in a web application called *EventEpi*. For each part, the IDB has to be preprocessed before any application of NLP, and texts of articles from RKI’s main online sources have to be extracted. The full pipeline is shown in Fig. 1. With the exception of the convolutional neural network for which we used Keras [7], we used the Python package scikit-learn to implement the machine learning algorithms [8].

**Figure 1:**
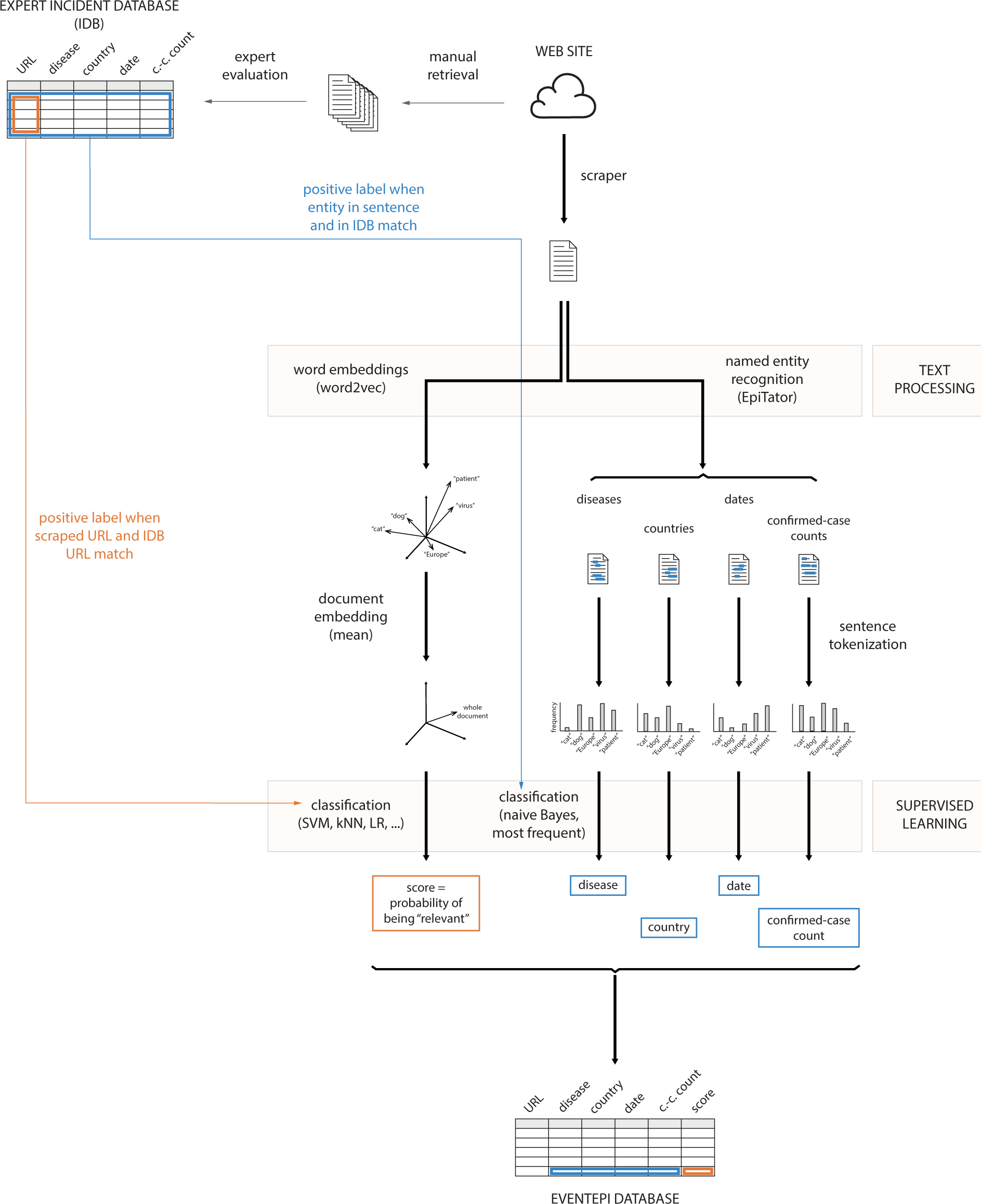
An illustration of the *EventEpi* architecture. The orange part of the plot describes the relevance scoring of epidemiological texts using document embeddings and different classification algorithms. The part of *EventEpi* that extracts the key information is colored in blue. Key information extraction is trained on sentences containing named entities using a naive Bayes classifier or the most-frequent approach. The workflow ends with the results being saved into the *EventEpi* database that is embedded into *EventEpi*’s web application.

### 2.1 Key information extraction

Key information extraction from epidemiological articles was in part already solved by EpiTator. EpiTator is a Python library to extract named entities that are particularly interesting in the field of epidemiology, namely: disease, location, date, and count entities. EpiTator uses spaCy^5^ (an NLP library) in the background to preprocess text. One function of EpiTator is to return all entities of an entity class (e.g., disease) found in a text. However INIG, as other EBS groups, is mostly interested in the *key* entities. Accordingly, the IDB contains a single value for each type of information. Thus, we needed to be able to condense the output of EpiTator to a single entity per class that best describes the corresponding event.

Besides other information, the IDB contains the mandatory columns **source, disease, country, confirmed-case count**, and the **date** of the number of confirmed cases of an outbreak article. To evaluate which of the entities found by EpiTator belong into the IDB, we compared the filtered EpiTator output of articles from the IDB (cf. Sec. 2.1.2) with the respective key information found in the IDB for this article. If the filtered output of EpiTator for a given article matched the key information columns of the IDB, then we knew that the filter selected the correct key entities.

To accomplish the key information extraction, two problems needed to be solved: First, the output of EpiTator needed to be comparable to the entries in the IDB. Second and more importantly, the output of EpiTator needed to be filtered.

#### 2.1.1 Preprocessing of the IDB

The IDB was not designed to be used with machine learning algorithms. It thus contained some inconsistencies that might not disturb human users but had to be resolved before machine learning algorithms were applied. This means that a column that would ask for a case count could also contain strings (of alphabetic letters) and a column that required a disease name could contain numbers. Other problems were that the entries in the IDB were written in German but the output of EpiTator is in English. Moreover, many entries were subject to typographic errors, formatting errors, and inconsistent naming schemes.

To translate German IDB entries into English, we built a translation dictionary. Since out of the mandatory columns, only the disease and country columns contained words, only two dictionaries were required. To translate the country names, we scraped the table found in the German Wikipedia article about the list of sovereign states (*Liste der Staaten der Erde*^6^) to obtain a mapping from several English and German variants of country names to the official English country name. For the translation of the disease names, we queried Wikidata^7^ for both German and English names of all diseases. Furthermore, we added disease abbreviations used internally at RKI as well as various intuitive custom abbreviations to the dictionary. As an example, the official abbreviation of the United Arab Emirates as found in the Wikipedia article is “ARE” or “AE”. However, more intuitive but unofficial abbreviations using the first capitals of the country or disease name, e.g., “UAE”, were often preferred in the IDB.

We used handwritten rules (e.g., removal of trailing whitespace) to correct for formatting errors. To maximize the number of IDB entries that are comparable to EpiTator’s output, we transformed the dataset into a tidy format [9] (e.g., spreading of several entries in one row to several rows). There are cases in which this approach led to an undesired outcome, e.g., when several countries are listed in a single row and the confirmed case count for this row is the sum of confirmed case counts of each listed country. Distributing the countries to several rows then leads to wrong case counts per row since the case count is just copied for each new entry. These cases are, however, seldom and the date and disease entires are still correct after spreading this row. If a word contained spelling mistakes, it could not be translated by our dictionaries. We searched for possible corrections of the word by minimizing the Levenshtein distance [10] of that word to all the words in our dictionary. The Levenshtein distance quantifies the amount of substitutions, additions, or deletions of characters to transform one word to another. We corrected a word if the number of corrections needed was lower or equal to 40% of the number of characters it contained. If no translation could be found, the word was kept unchanged.

#### 2.1.2 Key entity filtering

A naive approach to finding the key entity out of all the entities returned by EpiTator is to pick the most frequent one. We call this the *most-frequent approach*. This approach worked well for detecting the key country and disease, but not for the key date and confirmed-case count. For those, we developed a learning-based approach. This is shown in the *supervised learning* block in Fig. 1.

For the learning approach, we took the texts of the articles published in 2018 from the two most relevant sources, WHO DONs^8^ and ProMED^9^ (the reason for selecting those is described in Sec. 2.2.1) and applied a sentence tokenizer using the Python library NLTK [11]. (Tokenization is the process of splitting text into atomic units, typically sentences or words, including punctuation.)

We filtered all sentences to only keep those that contained some entity *e*_*c,j*_ recognized by EpiTator, with *c* being the class of the entity (date or confirmed-case count) and *j* being the *j*^*th*^ entity in a text. If an entity *e*_*c,j*_ in a sentence matched the entry of column/class *c* in the IDB, then we labeled this sentence as *key*. Every other sentence was labeled *not key*. We extracted samples from 3232 articles. The size of the datasets obtained is summarized in Tab. 1.

**Table 1:** Dataset sizes (number of sentences) for the key classes obtained from processing the articles in the IDB.

| key class | # positive samples | # negative samples | # total samples |
| --- | --- | --- | --- |
| confirmed case counts | 198 | 2245 | 2443 |
| date | 46 | 146 | 192 |

Then we trained a Bernoulli naive Bayes classifier (Bernoulli NBC) [12] with these labeled sentences to learn the relevant properties of sentences that led to the inclusion of their information into the IDB. Before applying a classifier, a text needs to be represented as a vector of numbers (vectorization). During training, a Bernoulli NBC classifier receives for each input sentence a binary vector *b* of the whole vocabulary (all the words seen during training) where the *i*^*th*^ position of the vector indicates the *i*^*th*^ term of the vocabulary. If the *i*^*th*^ term *t*_*i*_ is present in the input sentence, then *b*_*i*_ = 1 and 0 otherwise. The text is split into words using the NLTK word tokenizer. Additionally, we lower-cased all tokens which is standard procedure to avoid that tokens at the beginning of a sentence are treated differently. Finally, we applied stop word removal to reduce noise in our training data.

Based on the binary vectors and the corresponding labels, the Bernoulli NBC assigns probabilities to individual sentences of being *key* and *not key*. We decided that the key information for class *c* is the entity recognized in the one sentence, among all those containing an entity of class *c*, that has the highest probability of being *key*. This method assures that some entity is still chosen even though the classifier classifies no sentence as being *key*, i.e., all sentences in a text might have less than 50% probability of being *key*.

Additionally, we applied the multinomial NBC for comparison. The only difference to the Bernoulli NBC is that the multinomial NBC takes an occurrence vector *o*, with *o*_*i*_ being the frequency of term *t*_*i*_ in the text, as an input instead of a binary vector *b*. This approach is called bag-of-words. We combined bag-of-words with tf-idf (term frequency-inverse document frequency) where each term frequency is scaled so as to correct for overly frequent terms within and across documents. Formally, tf-idf is defined as

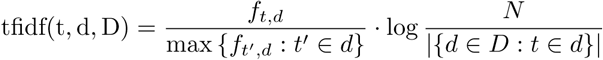

where *t* is a term from the bag-of-words, *d* is a document out of all epidemiological articles, *D* is the corpus of all epidemiological articles (containing *N* documents) and *f*_*t,d*_ is the frequency of term *t* occurring in document *d*. The Bernoulli NBC can be a better choice for short texts (such as single sentences), while the multinomial NBC often is preferred for longer text documents [12].

### 2.2 Relevance scoring

The second part of developing a framework to support EBS was to estimate the relevance of epidemiological articles. We framed the relevance evaluation as a classification problem, where articles that were present in the IDB are labeled *relevant*.

#### 2.2.1 Building the dataset

INIG uses a fixed set of sources and evaluates all articles from those sources. By scraping websites from INIG’s set of sources we could find all these articles, the *relevant* ones having the corresponding URL entered in the IDB, the rest being labeled as *irrelevant*. However it would have been too cumbersome to scrape all sources used in the IDB, as custom scrapers have to be built for each individual source. Therefore, we reduced the number of sources by firstly considering only sources that mostly contain epidemiological news (e.g., some websites contained mixed content of which only a fraction are outbreak news). Secondly, we chose sources that could be easily scraped. Thirdly, we picked those most frequently referenced in the IDB. Two sources stood out as being relevant, easy to scrape, and frequently entered in the IDB: World Health Organization Disease Outbreak News (WHO DON) and ProMED Mail. We had access to all assessments of the IDB of the year 2018 and therefore we scraped all WHO DON and ProMED articles of the year 2018. This resulted in a dataset of 3232 articles, 160 of them labeled *relevant* and 3072 *irrelevant*.

#### 2.2.2 Training of the classifiers

State-of-the-art text classifiers tend to use word embeddings [13, 14] for vectorization rather than the tf-idf and bag-of-words approach. Word embeddings are vector representations of words that are learned on large amounts of texts in an unsupervised-manner. Proximity in the word embedding space tends to correspond to semantic similarity. This is accomplished by assigning similar embeddings to words appearing in similar contexts. We applied word embeddings on lowercased words. First we used standard pre-trained embeddings, trained on the Wikipedia 2014 and Gigaword 5th Edition corpora^10^. However many terms, specific for epidemiology, were not represented. Thus, we produced custom 200-dimensional embeddings, training the word2vec [15] algorithm on the Wikipedia corpus of 2019^11^ and all available WHO DONs and ProMED Mail articles (more than 20,000 articles). We applied the skip-gram approach and hierachical softmax [15]. Those settings helped incorporating infrequent terms [16]. The embeddings were trained for five epochs.

Since we ultimately wanted to classify a whole document to find whether a text was *relevant*, we needed *document* embeddings. Although dedicated algorithms exist [17], we had not enough data to apply them meaningfully. However, taking the mean over all word embeddings of a document is a valid alternative [18] and suffices to show if learning the relevance of an article is possible.

A further issue was imbalance: Only a small fraction (5.0%) of the articles in the dataset was labeled *relevant*. Instead of discarding data from the majority class, we chose to up-sample the dataset using the ADASYN algorithm [19]. It generates new data points of the minority class by repeating the following steps until the proportion of minority and majority classes reaches the desired proportion (1:1):

1. choose a random data point *x*_*i*_ (the document embedding of article *i*) of the minority class;
2. choose randomly another minority-class data point *x*_*zi*_ among the 5-nearest neighbors of *x*_*i*_;
3. generate a new data point *y*_*j*_ at a random position between *x*_*i*_ and *x*_*zi*_ such that *y*_*j*_ = *a x*_*i*_ + (1*− a*) *x*_*zi*_ with *a* drawn uniformly at random between 0 and 1.

One problem of up-sampling data is that it still uses the minority class to create new examples and this might hinder the generalizability of the classifier [20].

Specific scores have been developed to gauge classification performance on imbalanced datasets. Here we consider the index balanced accuracy (IBA) [21]. It is defined as

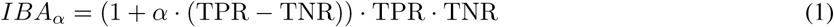

where 0 ≤ *α* ≤ 1 and *α* is a dominance index that needs to be fine-tuned based on how significant the dominating class is supposed to be. TPR and TNR are the true positive rate, i.e., the fraction of correctly classified relevant articles or key entities, and the true negative rate respectively. Following López et al., we use *α* = 0.1 and always report the weighted average of the IBAs where each class is respectively treated as the positive class. We used the imbalanced-learn package [22] to implement ADASYN and calculate the IBA.

We compared different classifiers for the relevance scoring task using embeddings or the bag-of-words approach:

- support-vector machine (SVM) using a penalty parameter of *C* = 1, kernel function coefficient 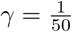, and a radial basis function as the kernel;
- k-nearest-neighbor classifier using *k* = 5, i.e., the five nearest neighbors, and the Euclidean distance to determine the neighborhood;
- logistic regression using L2 regularization of strength *C* = 1 to avoid overfitting;
- multinomial and complement [23] naive Bayes classifiers (NBCs), with priors equal to the class probability in the training data and as input, the tf-idf transformed bag-of-words with removed stop words;
- multilayer perceptron with 100 neurons in the hidden layer, with a rectified linear unit as their activation function, the Adam optimizer using the default values [24], and L2 penalty to avoid overfitting;
- convolutional neural network (CNN) using an singe convolutional layer with 100 filters, a kernel size of (1,2), stride of 1, ReLu activation function, max pooling over all filter, no padding, dropout of 0.25 followed by a fully connected layer with 256 hidden units and two output units using the softmax activation function; we used the Adam optimizer with an learning rate of 0.001.

SVM, k-nearest-neighbors, logistic regression and multilayer perceptron used document embeddings as features. The CNN operated on the word embeddings instead of the document embeddings. That way the striding filters of the CNN–if large enough–could learn relationships between adjacent words. We capped the input documents to a maximum of 200 words for the CNN. 89 documents contained less than 200 words which we filled up with zero embeddings such that each document has the same shape. For multinomial and complement NBCs, we used the bag-of-words approach since this feature representation coincides with the assumption of the NBC to predict a class given the occurrence (probability) of a feature. The features of word/document embeddings do not necessarily convey information about presence and absence of information. All classifiers received lowercased token and no punctuation.

The performances of all classifiers are evaluated on a test set which consists of 20% of the whole dataset. We applied stratification to assure that both classes are evenly distributed on the train and test set. The decision threshold for a class was set to 0.5, i.e., an input was attributed to the class of interest that had a probability above 0.5.

Finally, we used layer-wise relevance propagation [25] to make decisions of the CNN explainable. This is done by assessing which word embeddings passed through the CNN led to the final classification.

## 3 Results

In this section we present the performance of a series of key information extraction and relevance-scoring algorithms, and describe how the findings were embedded into the web application *EventEpi*.

### 3.1 Performance of key date and count extraction

We identified the most probable true entity among the many proposed by EpiTator using the most-frequent approach and two NBCs. Except for one occurrence of a disease that was not recognized by EpiTator, all countries and diseases were detected correctly using the most-frequent approach. However, this approach performed poorly for the key information extraction of date and case-count entities, with no date entity out of 141 (recall of 0 and IBA of 0 for the *key* class) and only 12 counts out of 105 (recall of 0.11 and IBA of 0.10 for the *key* class) correctly retrieved.

The performance of both NBC algorithms applied to extract key date and key confirmed-case count are shown in Tab. 2 and Tab. 3 respectively.

**Table 2:**
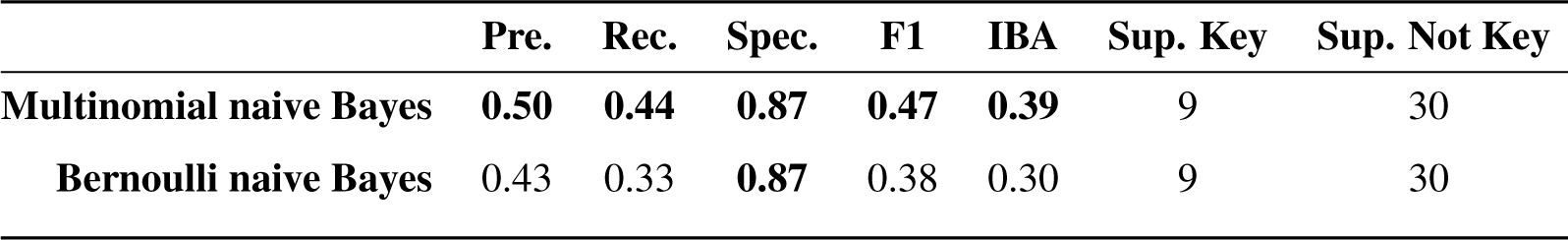
Evaluation of the *date* key information extraction. For each classifier and label, the precision (Pre.), recall (Rec.), specificity (Spec.), F1, index balanced accuracy (IBA) with *α* = 0.1, and support (Sup.) for both classes of the test set is given. The best values for each score highlighted in bold.

|  | Pre. | Rec. | Spec. | F1 | IBA | Sup. Key | Sup. Not Key |
| --- | --- | --- | --- | --- | --- | --- | --- |
| **Multinomial naive Bayes** | **0.50** | **0.44** | **0.87** | **0.47** | **0.39** | 9 | 30 |
| **Bernoulli naive Bayes** | 0.43 | 0.33 | **0.87** | 0.38 | 0.30 | 9 | 30 |

**Table 3:** Evaluation of the *confirmed-case count* key information extraction. Definitions and parameters are the same as in Tab. 2. The best values for each score highlighted in bold.

|  | Pre. | Rec. | Spec. | F1 | IBA | Sup. Key | Sup. Not Key |
| --- | --- | --- | --- | --- | --- | --- | --- |
| **Multinomial naive Bayes** | **0.32** | **0.65** | **0.88** | **0.43** | **0.58** | 40 | 449 |
| **Bernoulli naive Bayes** | 0.28 | 0.55 | **0.88** | 0.37 | 0.49 | 40 | 449 |

Besides IBA, we consider a number of scores defined as functions of TPR, TNR, the false positive rate FPR and the false negative rate FNR: precision = TPR*/*(TPR + FPR); recall also called sensitivity = TPR*/*(TPR + FNR); specificity = TNR*/*(TNR + FPR) and F1 = 2TPR*/*(2TPR + FPR + FNR). Since epidemiologists are interested in not missing any positives by classifying them incorrectly as negatives, we considered the recall as a good indicator for the performance of the classifiers. Because the dataset was imbalanced, we preferred the IBA as a measure for the overall accuracy.

For both date and count information extractions and all scores, multinomial NBC performed better or equal to Bernoulli NBC. Both Bernoulli and multinomial NBCs outperformed the most-frequent approach despite a small training dataset Tab. 1.

Thus, without offering perfect results, applying classification on named entity recognition as performed by EpiTator did improve key information extraction compared to the most-frequent approach. We expect these results to improve with time as data is accumulated and better structured.

### 3.2 Performance of relevance scoring

To compare the performance of the text classifiers, we looked at the same set of scores as for key information extraction. The results are shown in Tab. 4. While the SVM has the highest recall (0.88) and the CNN has the highest precision (1.00), both classifiers seem to overfit one class. The MLP yields the best trade-off between the different metrics, having the highest F1 score (0.28) and the highest IBA (0.46), while still having a fairly good recall (0.5).

**Table 4:**
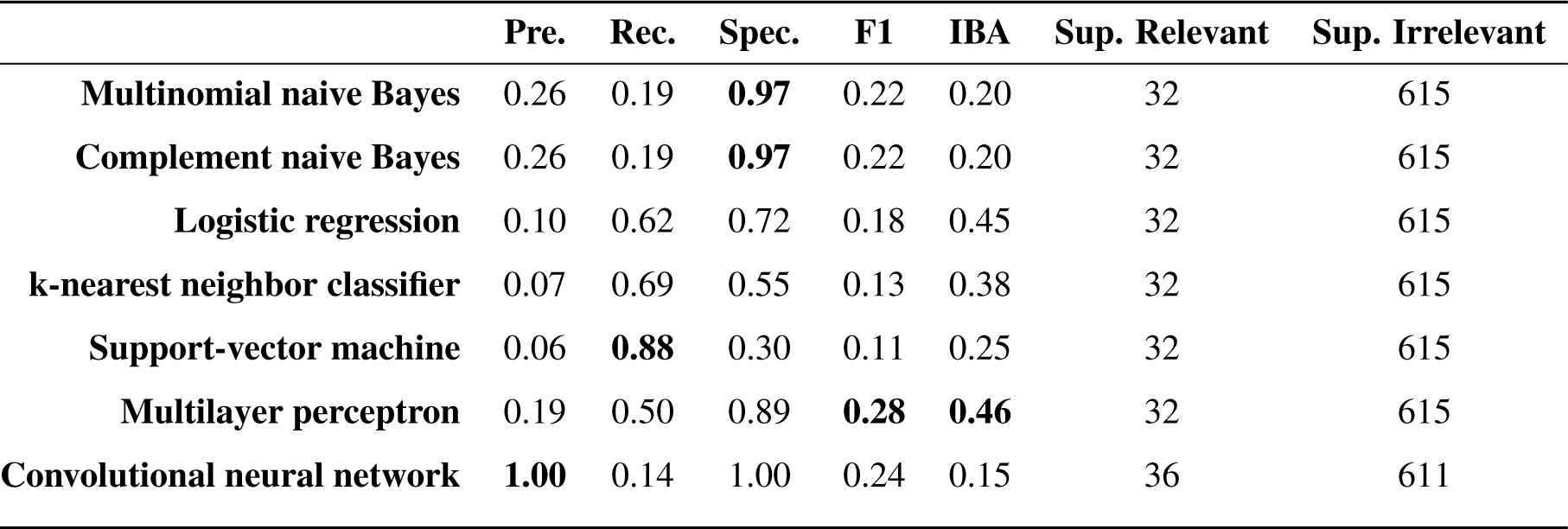
The performance evaluation of the relevance classification. For each classifier and label, the precision (Pre.), recall (Rec.), specificity (Spec.), F1, index balanced accuracy (IBA) with *α* = 0.1, and support (Sup.) for both classes of the test set is given. The best values for each score are highlighted in bold.

Against expectations, the multinomial and complement NBCs had an identical performance although the complement NBC tackles problems occurring in imbalanced datasets specifically. A reason for the poor performance of the CNN is that it overfitted to the majority class, classifying almost all articles as *irrelevant*. Overfitting generally can be avoided with further dropout (random removal of nodes in the network during training time to minimize highly specified nodes), regularization (e.g., L2 to punish strong weighting of nodes), and early stopping (to minimize the difference of losses between the test and validation set). However, the data set is probably too small for CNNs to yield significant improvements. Note that the dataset for CNN was sampled in a seperate process which led to a test set of same size but a slightly higher number of *relevant* articles.

It is nevertheless interesting to use the CNN as an example for explaining what contributed to the classification. A plot of a layer-wise relevance propagation shows one example where the a relevant article was correctly classified as being *relevant* (Fig. 2). We see that words like *500* in the beginning of the text are highlighted as strongly important for the classification of the text as being *relevant*. Also, the word *schistosomiasis*–an infectious disease caused by flatworms–is labeled as strongly relevant for the classification. Interestingly, it is also relevant for the classifier, that this disease is treated with antiparasitic drugs (*anthelmintic*). Both makes sense, since a very high number of cases of a dangerous infectious disease are of interest for epidemiologists. All other case numbers are labeled as slightly irrelevant which does not necessarily make sense. An event might be less relevant when out of 500 confirmed cases of some infectious disease half of the patients are in treatment.

**Figure 2:**
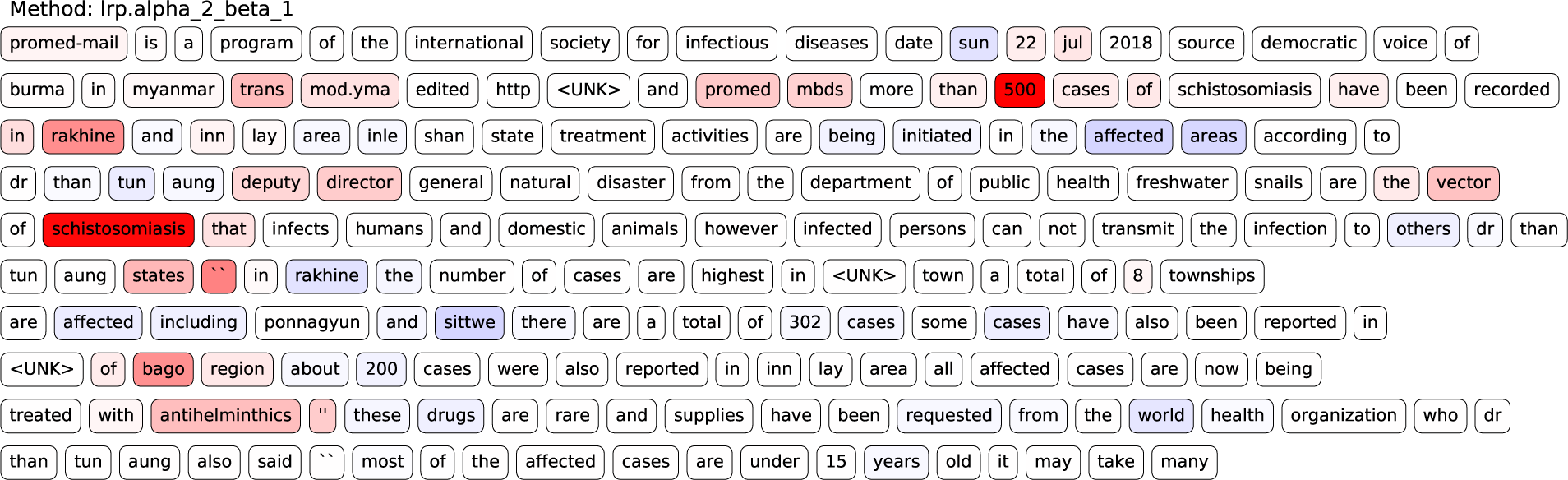
A layer-wise relevance propagation of the CNN for relevance classification. This text was correctly classified as relevant. Words that are highlighted in red contributed to the classification of the article being *relevant* and blue words contradicted this classification. The saturation of the color indicates the strength of which the single words contributed to the classification. *<UNK>* indicates a token for which no word embedding is available.

The focus of this work was to show a proof of concept that classification methods can serve in determining the relevance of an article. We did not try to fine-tune all of the compared classifiers. For now, SVM, although having a low precision, is preferred due to its good recall, i.e., a low chance to miss true positives during EBS. The multilayer perceptron is also a good choice due to its high IBA. Although the relevance classification has not a strong performance, it could already aid epidemiologists. The model could be retrained every time articles are entered into the IDB to increase performance continuously. Until then, the relevance score could be displayed and used to sort articles, but not to filter content.

### 3.3 Web service

To showcase the analyses presented above and show how key information and relevance scoring can be used simultaneously to aid EBS, we developed the web application *EventEpi*. Fig. 3 shows a screenshot of its user interface. *EventEpi* is a Flask^12^ app that uses DataTables^13^ as an interface to its database. *EventEpi* lets users paste URLs and automatically analyze texts from sources they trust or are interested in. The last step in Fig. 1 shows how the *EventEpi* database is filled with the output of the key information extraction and relevance scoring algorithms. With our colleagues at INIG in mind, we integrated a mechanism that would automatically download and analyze the newest unseen articles from WHO DONs and ProMED. Currently, this process is slow and depends on pre-analyses for a good user experience. To allow for the integration of the functionality into another application, we also wrote an ASP.NET Core application that allows to analyze texts via an API calls.

**Figure 3:**
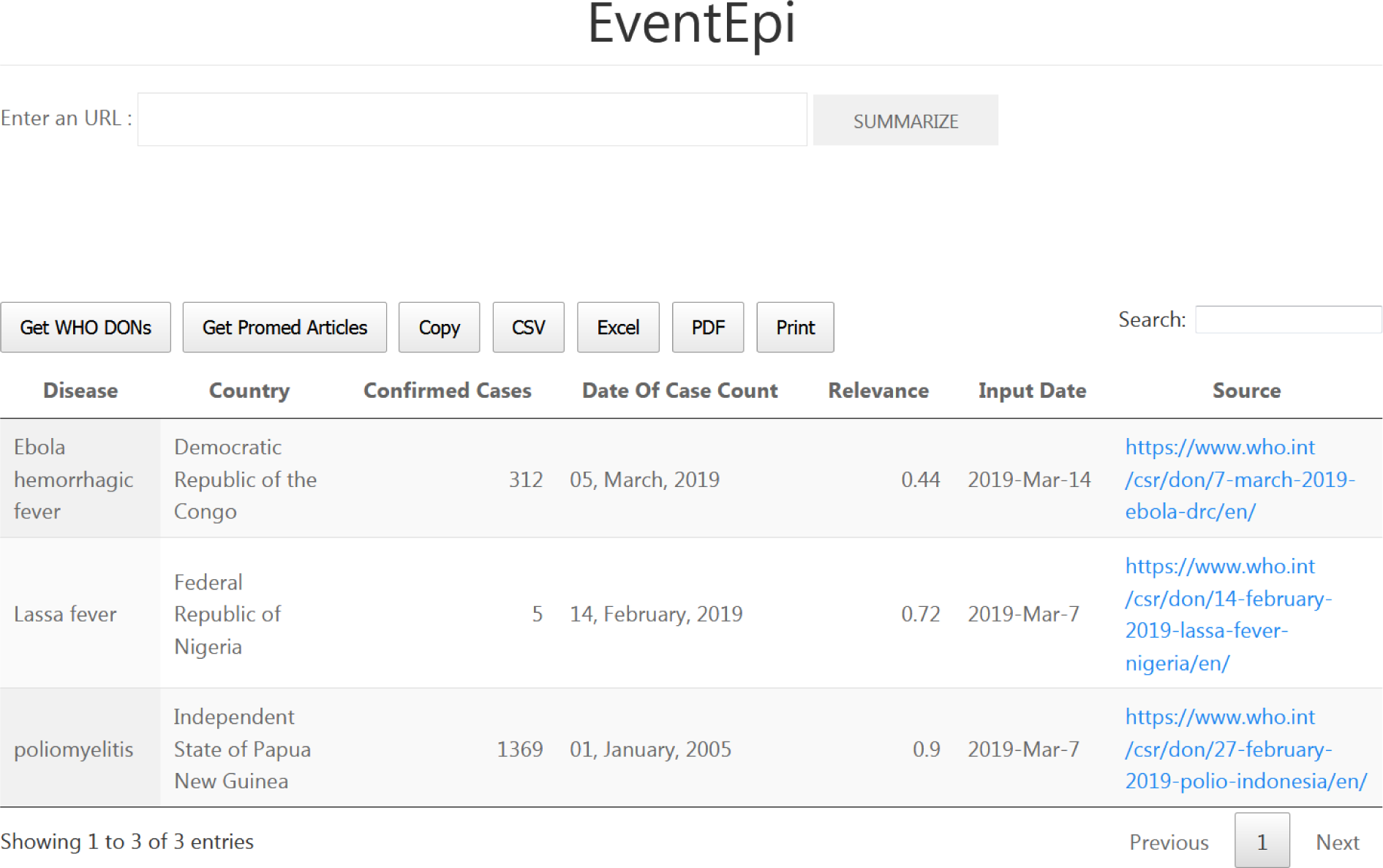
A screenshot of the *EventEpi* web application. The top input text field receives an URL. This URL is summarized if the `SUMMARIZE` button is pushed. The result of this summary is the input into the datatable which can be seen in the center of the image. The buttons `Get WHO DONs` and `Get Fromed Articles` automatically scrape the last articles form both platforms that are not yet in the datatable. Furthermore, the user can search for words in the search text field and download the datatable as CSV, Excel or PDF.

## 4 Conclusion

We have shown that novel NLP methodology can be applied in combination with available resources, in this case the IDB of the RKI, to improve epidemiological surveillance. Even with limited datasets, EBS can be supported by automatic processes, and in the future even partially automated.

More work is necessary to bring *EventEpi* into production. While key disease and country can satisfactorily be extracted, the performance of key date and confirmed-case count extractions needs to be improved. The moderate performance was presumably mostly due to the small number of labeled data.

Relevance scoring shows promising results. We believe it could already be helpful to epidemiologists, and could greatly be improved with fine-tuning and larger datasets.

The web application *EventEpi* is a scalable tool. Thus, the scope of EBS might be increased without comparable increase in effort. This is particularly relevant with the availability of automatic translation (for example DeepL^14^). It could allow an EBS team to access much more sources than those in the few languages its members typically speak without being overwhelmed. It is possible to provide better classifications that work for different languages using multilingual word embeddings [26], or a better key information extraction using contextual embeddings [27, 28] which adjust the embedding based on the textual context. Contrary to the relevance of a document, key information is mostly defined by its nearby words.

The same fundamental issues encountered in using machine learning in general apply here as well, in particular bias and explainability. Tackling individual biases and personal preferences during labeling by experts is essential to continue this project and make it safe to use. It will also be important to show *why EventEpi* extracted certain information or computed a relevance, for it to be adopted but also critically assessed by epidemiologists for improvement. For artificial neural networks, we showed that layer-wise relevance propagation can be used in the domain of epidemiological texts to make a classifier explainable. For other models, model agnostic methods [29, 30] could be applied analogously.

At the moment *EventEpi* only presents results to the user. However it could be expanded to be a general interface to an event database and allow epidemiologists to note which articles were indeed relevant as well as correct key information. This process would allow more people to label articles and thus expand the data sets, as well as help better train the relevance-scoring algorithms, an approach called active-learning [31].

With a large labeled dataset, a neural network could be (re)trained for the relevance classification. Later, transfer learning (tuning of the last layer of the network) could be used to adapt the relevance classification to single user preferences.

## Data Availability

The data is not available. The source code is publicly available.

https://github.com/aauss/EventEpi

## 5 Acknowledgements

We would like to thank Maria an der Heiden, Sandra Beermann, Sarah Esquevin, Raskit Lachmann and Nadine Zeitlmann for helping us on questions regarding epidemiology, providing us with data and for critical comments on the manuscript. We also kindly thank Katarina Birghan for helping us using Wikidata and Fabian Eckelmann for his support on developing the *EventEpi* web application.

## Footnotes

1 https://grits.eha.io

2 https://github.com/ecohealthalliance/EpiTator

3 http://medisys.newsbrief.eu/medisys/helsinkiedition/en/home.html

4 http://puls.cs.helsinki.fi/static/index.html

5 https://spacy.io/

6 https://de.wikipedia.org/wiki/Liste_der_Staaten_der_Erde

7 https://w.wiki/4wJ

8 https://www.who.int/csr/don/en/

9 https://www.promedmail.org/

10 https://www.kaggle.com/rtatman/glove-global-vectors-for-word-representation

11 https://dumps.wikimedia.org/

12 http://flask.pocoo.org/

13 https://datatables.net/

14 https://www.deepl.com/translator

